# Automatic colorectal cancer screening using deep-learning on spatial light interference microscopy data

**DOI:** 10.1101/2022.01.16.22269381

**Authors:** Jingfang K. Zhang, Michael Fanous, Nahil Sobh, Andre Balla, Gabriel Popescu

## Abstract

The surgical pathology workflow currently adopted in the clinic uses staining to reveal tissue architecture within thin sections. A trained pathologist then conducts a visual examination of these slices and, as the investigation is based on an empirical assessment, a certain amount of subjectivity is unavoidable. Furthermore, the reliance on such external contrast agents like hematoxylin and eosin (H&E), albeit a well-established method, makes it difficult to standardize color balance, staining strength, and imaging conditions, hindering automated computational analysis. In response to these challenges, we applied spatial light interference microscopy (SLIM), a label-free method that generates contrast based on the intrinsic tissue refractive index signatures. Thus, we reduce human bias and make the image data comparable across instruments and clinics. We applied a Mask R-CNN deep learning algorithm to the SLIM data to achieve an automated colorectal cancer screening procedure, i.e., classifying normal vs. cancer specimens. Our results obtained on a tissue microarray consisting of specimens from 132 patients, resulted in 91% accuracy for gland detection, 99.71% accuracy in gland-level classification, and 97% accuracy in core-level classification. A SLIM tissue scanner accompanied by an application-specific deep learning algorithm may become a valuable clinical tool, enabling faster and more accurate assessment by the pathologist.

## INTRODUCTION

From benign adenomatous polyps to carcinoma, colorectal cancer develops through a series of genetic mutations over a 5-10 year course [1]. The mortality of colorectal cancer is significantly reduced if this disease can be diagnosed at an early, “localized” stage. The 5-year survival rate for patients with their disease diagnosed at the localized stage is 89.8%, while the same rate for patients with distant metastasis (late-stage cancer) drops to 12.9% [2]. In the United States, the preferred form of screening is colonoscopy. In the 50-75 age group, 65% of individuals received colorectal cancer screening in 2010, compared to 54% in 2002 [3]. As colorectal cancer has become the third most prevalent cancer diagnosed in the U.S., the American Cancer Society estimates that there will be 149,500 new cases in 2021 and 52,980 associated deaths [4]. The U.S. Preventive Services Task Force (USPSTF) recommended in 2016 [5] that all 50-75 year old adults receive colorectal cancer screening, but then in 2020, it issued a revised draft, which notably decreased the starting age for all-adults screening to 45-49 [6]. Among all the individuals receiving colonoscopy, the prevalence of adenoma is 25-27%, and the prevalence of high-grade dysplasia and colorectal cancer is just 1-3.3% [7, 8]. However, 50% of all colonoscopies underwent a polyp removal or biopsy as current screening tools cannot effectively distinguish adenoma from a benign polyp [9]. Following the procedure, pathologists examine the excised polyps to identify whether the tissue is cancerous or benign. As more individuals are expected to receive colonoscopy in the future, pathologists will likely see a significantly heavier workload. For the purposes of large-scale efficient screening, it is imperative to acquire new technologies that can decrease manual work by performing of automated tissue investigation. The Papanicolaou test (pap smear) for cervical cancer screening is a successful precedent, which employs semi-automated computational tools and, in the example of FDA-approved BD Focal Point Slide Profiler™, decreases pathology caseloads by 25% by detecting benign cases [10]. It must be noted, however, that a proper operation of such systems requires staining procedures tailored for narrow calibration thresholds, and continuous quality-reviews are required throughout the machine’s operation [11].

Quantitative phase imaging (QPI) [12] has been established as a valuable *label-free* method for various biomedical imaging applications [13]. Due its sensitivity to tissue nanoarchitecture and ability to provide quantitative information from 3D and anisotropic tissue structures [14-16], QPI has been applied to different pathology problems in recent years [12, 17-24]. Spatial light interference microscopy (SLIM) [25-27] has been used as the core technology for label-free whole slide imaging (WSI), which revealed that tissue refractive index is an effective intrinsic marker for pathological diagnosis and prognosis [21, 22, 28-34]. It was shown that tissue scattering coefficients computed from the QPI data were able to predict disease recurrence after prostatectomy surgery [20, 21]. Furthermore, it has been demonstrated that SLIM could provide information on collagen organization, which represents valuable information for disease stratification in breast cancer pathology [22, 30, 35].

Although this approach of “feature engineering” has the advantage of generating physically insightful markers [36] (e.g., scattering anisotropy factor, refractive index variance), it encompasses a limited range of the parameters that can be retrieved from these data [37]. There is always the possibility that certain clinically valuable parameters remain unevaluated. In recent years, the biomedical community has applied artificial intelligence (AI) techniques to data processing, visualization, and analysis [37-45]. Contrary to feature engineering, a deep learning convolutional neural network extracts a large number (millions) of features, including edges, pixel intensities, variations in pixel values, etc., for each image. AI can recognize image patterns that might be too subtle for human eyes, and thus significantly improve clinical cancer screening and diagnosis. In this scientific area, we recently have demonstrated that the combination of SLIM and AI can screen colorectal tissue as cancer vs. benign [37]. However, while very encouraging, this previous work required segmented individual glands as input. In other words, a prerequisite for this procedure was the manual annotation of gland regions, which diminished the practical use of tour method.

In order to advance our screening method and provide completely automatic classification, here we used a SLIM-based whole slide imaging tissue scanner together with a Mask R-CNN deep learning network to both segment the glands and classify cancer and benign tissues. This new method of automatic colorectal cancer screening uses intrinsic tissue markers and, thus, is now of potential clinical value. Importantly, this method does not require staining or calibration. Unlike the existing staining markers, the signatures that are derived from phase information can be shared across different instruments and laboratories, without modification. Furthermore, we eliminated the need for manual segmentation as a pre-requisite of the procedure.

## RESULTS AND METHODS

### Label-free tissue scanner

Our label-free WSI system consists of a SLIM scanner, with specially designed hardware and software [11]. As shown by Figure 1, the SLIM module (Cell Vista SLIM Pro, Phi Optics, Inc.) is an upgrade module to an existing phase-contrast microscope [25, 26], which generates quantitative phase maps associated with the sample. Fundamentally, SLIM performs by making the ring in phase contrast objective pupil “tunable”. To achieve this, the image generated by a phase contrast microscope is Fourier transformed at a spatial light modulator (SLM) plane, where the image of the phase contrast objective ring is overlaid with the SLM phase mask, and the phase steps are shifted incrementally by 90° (Fig 1). From the four intensity images associated with the quarter wavelength phase shifts, the quantitative phase image is extracted to reveal the optical pathlength at each point in the field of view.

**Figure 1:**
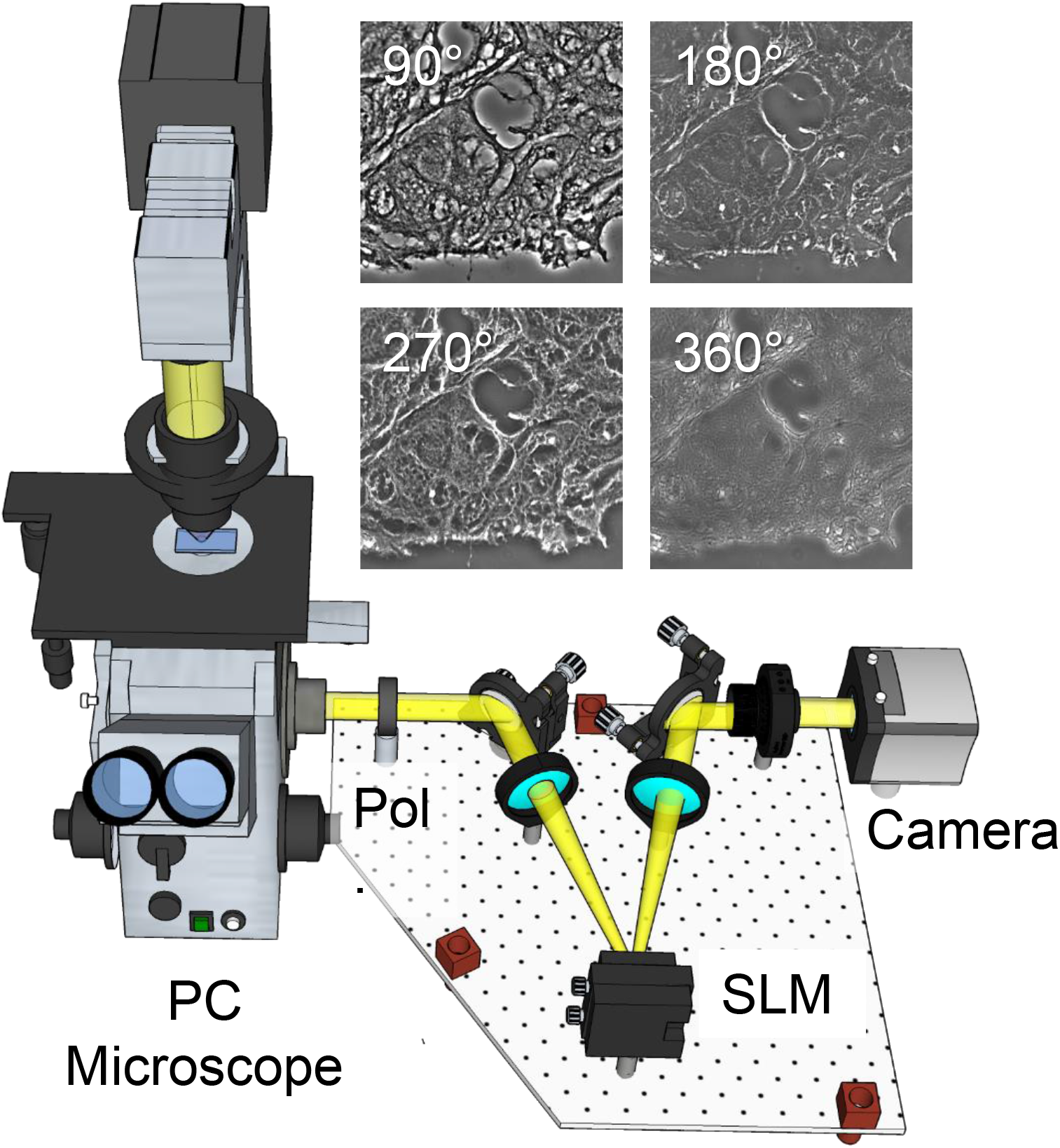
SLIM tissue scanner setup. SLIM system implemented as add-on to an existing phase contrast microscope. Pol, polarizer and SLM, spatial light modulator. The four independent frames corresponding to the 4 phase shifts imparted by the SLM are shown for a tissue sample.

Equipped with a novel acquisition software that pairs up both CPU (central processing unit) and GPU (graphical processing unit) processing, the SLIM tissue scanner can acquire and process the four intensity images and display the phase image in real time [11]. The SLIM phase retrieval computation is performed at a different thread, while the microscope stage shifts the subsequent position. The scanning of a large field of view like a whole microscope slide and the assembling of the ensuing images into a single file are performed by our specifically designed software tools [11]. The final SLIM images are generated at maximum 15 frames per second, as restricted by the refresh rate of the spatial light modulator.

### Tissue imaging

We used archival pathology material from 131 patients who went to the University of Illinois at Chicago (UIC) in 1993-1999 to receive colorectal resection for cancer treatment. Colon tissue cores of 0.6 mm diameter were collected from each patient, which corresponded to four groups: “tumor”, “normal mucosa”, “dysplastic mucosa”, and “hyper plastic mucosa”[11]. The tissue cores were finally transferred into high-density arrays for the purpose of imaging: primary colon cancer (127 cases), mucosa of normal colon (131 cases), dysplastic colon (33 cases), and hyperplastic colon (86 cases) [11].

From each specimen, two 4-μm thick sections were cut. The first section was deparaffinized and stained with hematoxylin and eosin (H&E) and imaged using the Nanozoomer (bright-field slide scanner, Hamamatsu Corporation). A pathologist made a diagnosis for all tissue cores in the TMA set, which was used as “ground truth” for our analysis. A second adjacent section was prepared in a similar way but without the staining step. These slides were imaged in our laboratory. All these studies complied with the protocols that the Institutional Review Board at the University of Illinois at Urbana-Champaign approved (see IRB Protocol No. 13900). Before the imaging, the tissue slices were deparaffinized and cover-slipped in aqueous mounting media. A conventional microscope (Zeiss Axio Observer, 40×/0.75, AxioCam MRm 1.4 MP CCD) acquired a series of mosaic tiles of the tissues, which were then assembled into tissue microarray images. With ImageJ’s stitching functionality, we tiled high-resolution images of each core. Using Cell Vista SLIM Pro (Phi Optics, Inc.) 1.2 × 1.2 mm^2^ regions, consisting of 4 × 4 mosaic tiles, were SLIM-imaged at a 0.4 μm resolution, in four seconds. For each frame, stage motion takes 100 ms, SLM modulator stabilization 30 ms, and exposure 10 ms. From the resulting image file, 10,000*10,000 pixels images were cropped, corresponding to a field of view at 1,587.3 × 1,587.3 μm^2^.

For this work, only tumor and normal mucosa images were used, since dysplastic mucosa (33 samples) and hyper plastic mucosa (86 samples) images were not enough to support our training. From the 127 primary colon cancer images and the 131 normal colon mucosa images, a total of 258 images were selected. Of them, 98 cancer images and 98 normal images were assigned for the training, 15 cancer images and 15 normal images assigned for the validation, and 14 cancer images and 18 normal images assigned for the testing. As a result, 76% of the dataset are used for training, 12% for validation, and the remaining 12% for testing. The 32 testing images were hidden until all the training and fine-tuning was completed and finalized.

### Deep learning network

We used deep learning for the semantic segmentation of tissue glands. For this purpose, three steps are required: (1) object recognition; (2) object localization; and (3) object classification [46]. In object detection, a bounding box with (x, y) coordinates together with a class label are predicted for each object. In semantic segmentation, the input image is partioned and every pixel value is predicted and assigned to a certain class label. In addition to the semantic segmentation, instance segmentation adds a pixel-wise mask to each object detected.

A Mask R-CNN deep learning network was set up with an algorithm that He et al. published in 2017 [47, 48], which is a further improvement on the previous R-CNN, Fast R-CNN[49] and Faster R-CNN [50] algorithms. On top of the Faster R-CNN architecture that predicts class labels and bounding boxes, Mask R-CNN adds a two-CONV-layers branch (Fig 2) that predicts the mask for each detected object and gives a pixel-wise visualization of each object’s location. Our Mask R-CNN network [48] uses Keras and TensorFlow and loads a pretrained ResNet101 network from the ImageNet database. The residual connections in this algorithm can improve gradient flow and, thus, boost the training of deeper networks. With our dataset of 196 images for training and 30 images for validation, ResNet101, with its 101 layers, extracted valuable features and supported a high accuracy in gland detection and classification.

**Figure 2:**
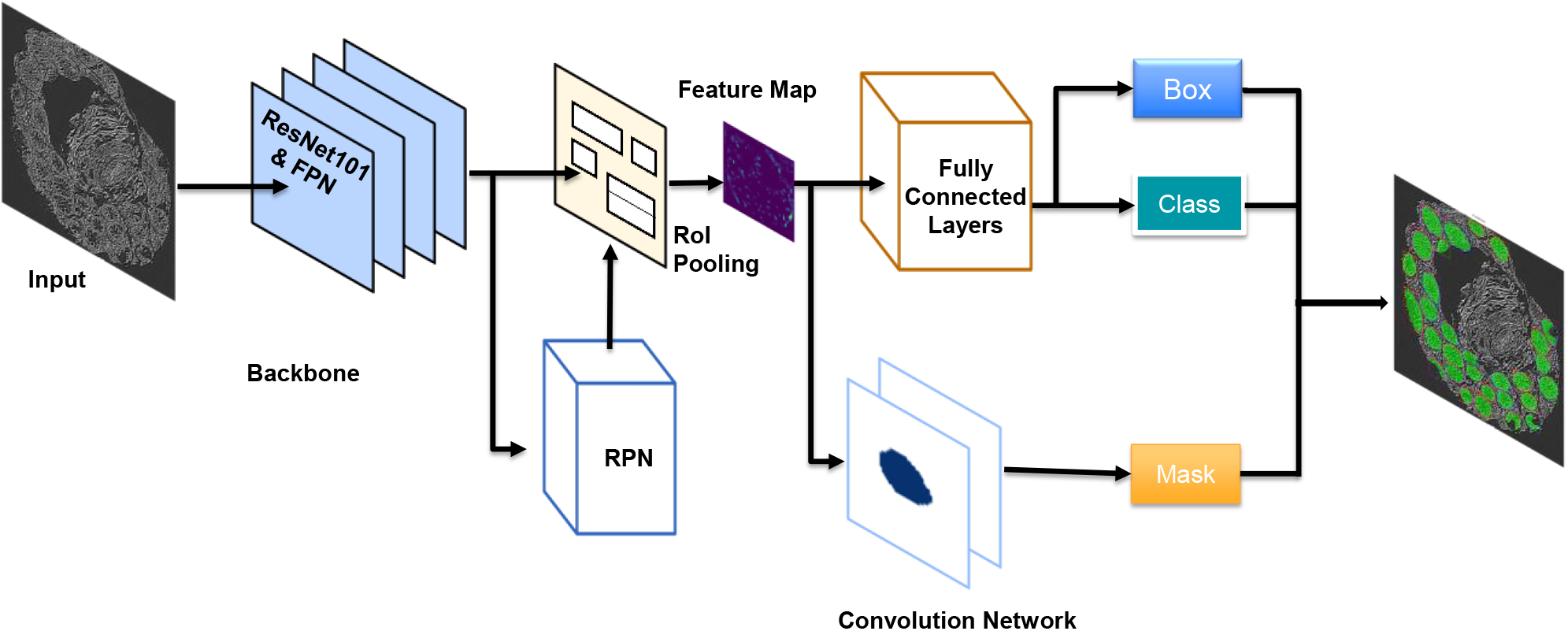
Mask R-CNN network architecture. Mask R-CNN (regional convolutional neural network) framework contains two stages: scan image and generate regional proposals for possible objects; classify the proposals and generate bounding boxes and pixel-wise masks. This specific network adopts a backbone of ResNet101 plus FPN for feature extraction. RPN (Region Proposal Network) scans over backbone feature maps, which allows reuse of the extracted features and remove duplicate calculations. RPN outputs two results for each anchor: anchor class (Foreground or Background: Foreground implies potential existence of an object) and bounding box refinement (The foreground anchor box, with its location and size, is refined to fit the object). The final proposals are passed to the next stage. At this stage, two outputs are generated for each ROI as proposed by the RPN: class (for objects), and bounding box refinement. ROI Pooling algorithm crops a piece of area from a feature map and resizes it to a fixed size, which enables the functionality of classifiers. From this stage, a parallel branch, of two fully convolution layers, is added that generates masks for the positive regions that are selected by ROI classifier. The other branch, of fully connected layers, takes the outputs of the ROI Pooling and outputs two values: a class label and a bounding box prediction per object.

In our Mask R-CNN network there were three classes: “cancer”, “normal” and “background”, in which only “cancer” and “normal” were flagged. We trained the network using a single GPU (with a 12 GB memory) and set the batch size at 1. The RPN (Region Proposal Network) adopts (8, 16, 32, 64, 128) as anchor box scales and [0.5, 1, 2] as anchor ratios. 8, 16, 32, 64 and 128 are respectively the length of square anchor side in pixels, which means that 5 square anchor boxes of 8*8, 16*16, 32*32, 64*64 and 128*128 pixels are generated for each anchor point. With the three anchor ratios, a total of 5*3 anchor boxes is generated for each anchor point. We believed that such a combination of our anchor scales and anchor ratios would ensure a satisfying performance in our gland detection purpose. Anchor scales should be adjusted as per the overall shapes of the objects that are detected. Full-size masks are used throughout our training process, which means the masks are set to have the same width and height of the original image. Another alternative method is to use mini masks during the training, which can be generated by extracting the bounding box of the object and resize it to a pre-defined shape. For example, a full-size mask of 1024*1024 pixel can be resized to a size of 224*224 pixel, which reduces memory requirement during trainings. However, mini masks likely do not facilitate training performance in case of small-size objects, such as cells, since minor rounding errors may arise while the masks are converted to a smaller size and then eventually converted back. The optimizer adopts ADAM, which is combined with Adaptive Gradient Algorithm (AdaGrad) and Root Mean Square Propagation (RMSProp). AdaGrad improves performance in cases with sparse gradients, while RMSProp algorithm works well in cases with noise. Learning rate is 0.001, learning momentum 0.9 and weight decay 0.0001. A moderate level of image augmentation is implemented, which includes horizontal flipping 50% of all images, vertical flipping 50% of all images, affine rotate by 90°, 180° and 270°, and gaussian blur with 0.0-5.0 sigma.

The machine in this work has 1 GPU, NVIDIA RTX 2070. It took 46 hours 30 minutes to complete 400 epochs of training. The first 50 epochs trained only top layers and the remaining 350 epochs transferred to train all the 101 layers. The images were annotated with VGG Image Annotation tool, and data was written into JSON file. The gland detection confidence probability was tested separately at 0.70, 0.80 and 0.90, from which 0.90 was selected for the final generalization. All the testing statistics in this paper are based on the 390^th^ epoch and the 0.90 detection confidence score, unless noted otherwise.

### Whole core segmentation and classification

As normal glands tend to show clear and regular edges, they can be predicted and classified with higher accuracy. Cancer glands tend to show irregular and less pronounced edges, which makes it more difficult for the network to predict cancer glands. Figure 3 exemplified the results of gland segmentation for both normal and cancer glands. Figure 3b is a cancer core used as input to the network, while Fig. 3c is the output of the network, which displays segmentation, classification, and masking information. Red masks represent cancer glands. Figures 3e-f represent the analog pair for a normal core, where green masks represent normal glands. Figures 3d-g display individual glands from Fig. 3c and 3f, respectively, as indicated.

**Figure 3:**
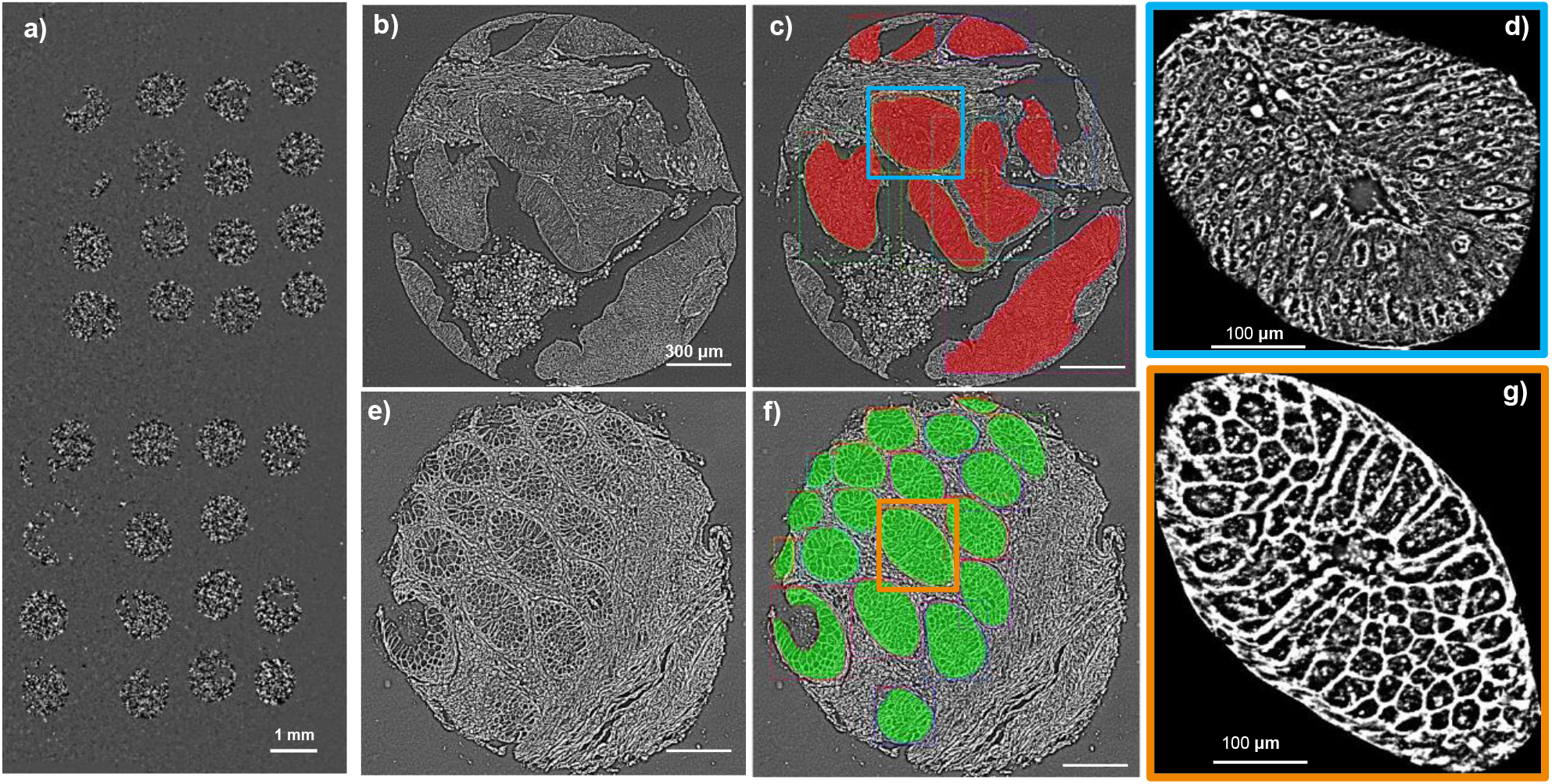
Examples of Segmentation and Classification. a) Images of 32 testing cores. b) Cancer core. c) Prediction of gland detection and classification on the core in b). d) Zoom-in image of the cancer gland boxed in c). e) Normal core. f) Prediction of gland detection and classification. g) Zoom-in image of the normal gland boxed in f). Red color represents cancer and green color normal glands.

### Overall gland classification and detection

Figure 4 provides four cases of segmentation output, displaying different performances for *cancer* gland detection and *normal* gland detection. The network segments with higher accuracy normal than cancer glands. In Fig. 4a the network failed to detect two cancer gland areas, which were highlighted by white boxes in Fig. 4b. In Fig. 4c, two cancer gland areas are also missed, as highlighted by the white boxes in Fig. 4d. In Figs. 4e, g, all the glands are correctly detected and classified, which can be seen Figs. 4f, h. In addition, two more true normal glands, which were not annotated in the 4e ground truth core and appeared tattered, are detected and classified correctly in 4f; and three such cases in 4g-h.

**Figure 4:**
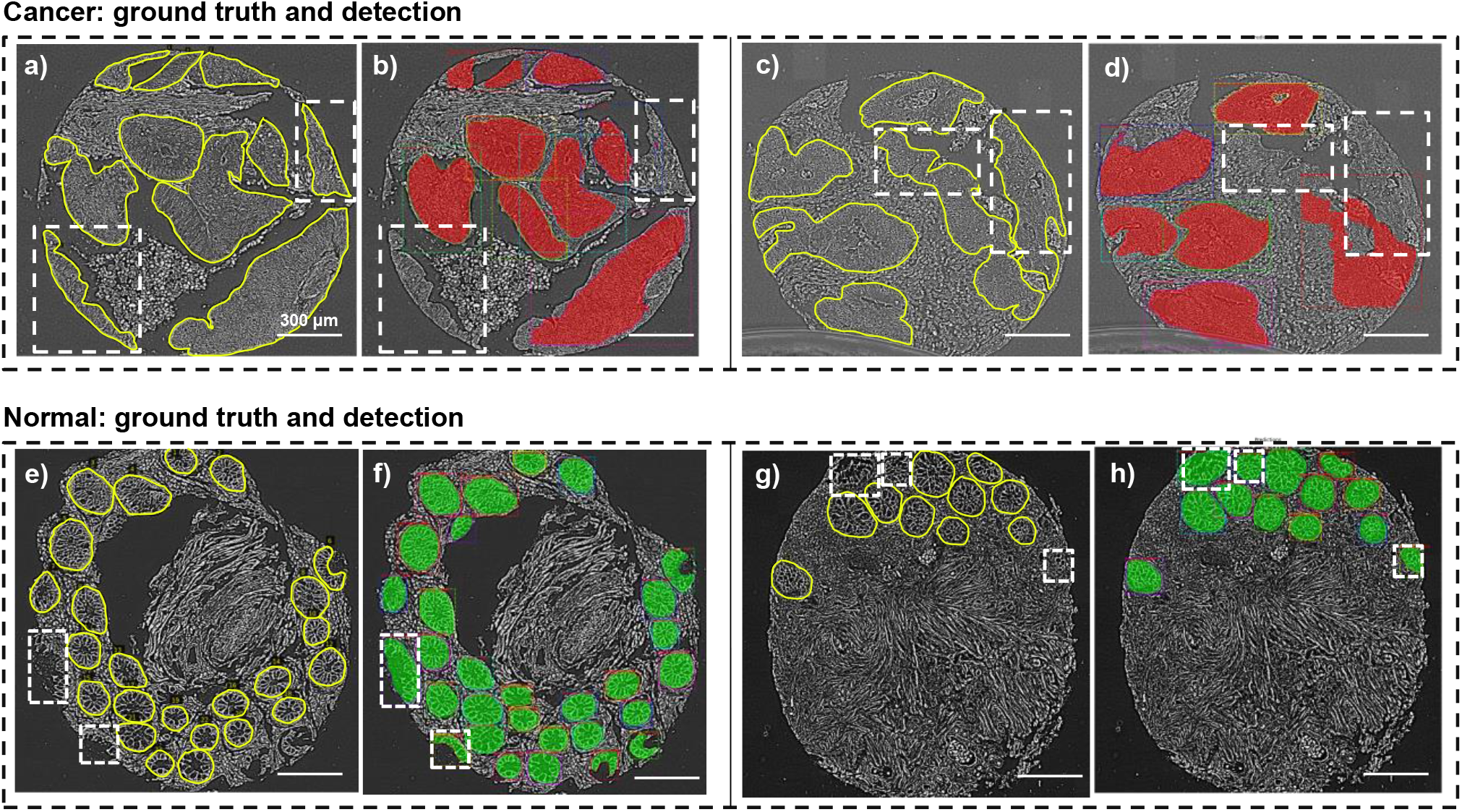
Examples of Gland Detection Errors and Additional Positive Detections. a) Ground truth (manual) segmentation of cancer glands. b) Network prediction, showing that the regions in the dash boxes were missed. c-d) Similar illustration as in a-b). e) Ground truth (manual) segmentation of normal glands. f) Network prediction, showing that the regions in the dash boxes were additional true positives. g-h) Similar illustration as in e-f). Note that all errors and additons occur at the boundaries of the cell cores.

Figure 5a and 5b show overall performance in gland classification and gland detection by means of confusion matrix. In Fig. 5a, the first row represents the actual class of cancer glands, where 95 instances are correctly classified and 1 instance is wrongly classified as normal. Second row represents the actual class of normal gland, where all the 248 normal glands that are captured are correctly classified. In Fig. 5b, the first row represents the actual class of cancer gland. Of the total of 116 ground truth cancer glands, 96 cancer glands are detected, of which 1 of the 96 instances misclassified as a normal gland and the remaining 95 cancer glands correctly classified. As a total, 20 cancer glands are not detected, which are misclassified as stroma or background. The second row represents the actual class of normal gland, of which 248 of the total 251 normal glands are detected and correctly classified and 3 normal glands are not detected. The third row represents stroma or background, where 5 stroma tissues are detected and classified as cancer gland while 4 stroma tissues are detected and classified as normal gland. Of all the ground truth cancer glands, 83% are detected and 17% missed. Of all the ground truth normal glands, 99% are detected and 1% missed. Of all the detected instances, 2% are non-gland instances. Of all the ground truth glands, 6% are not detected.

**Figure 5:**
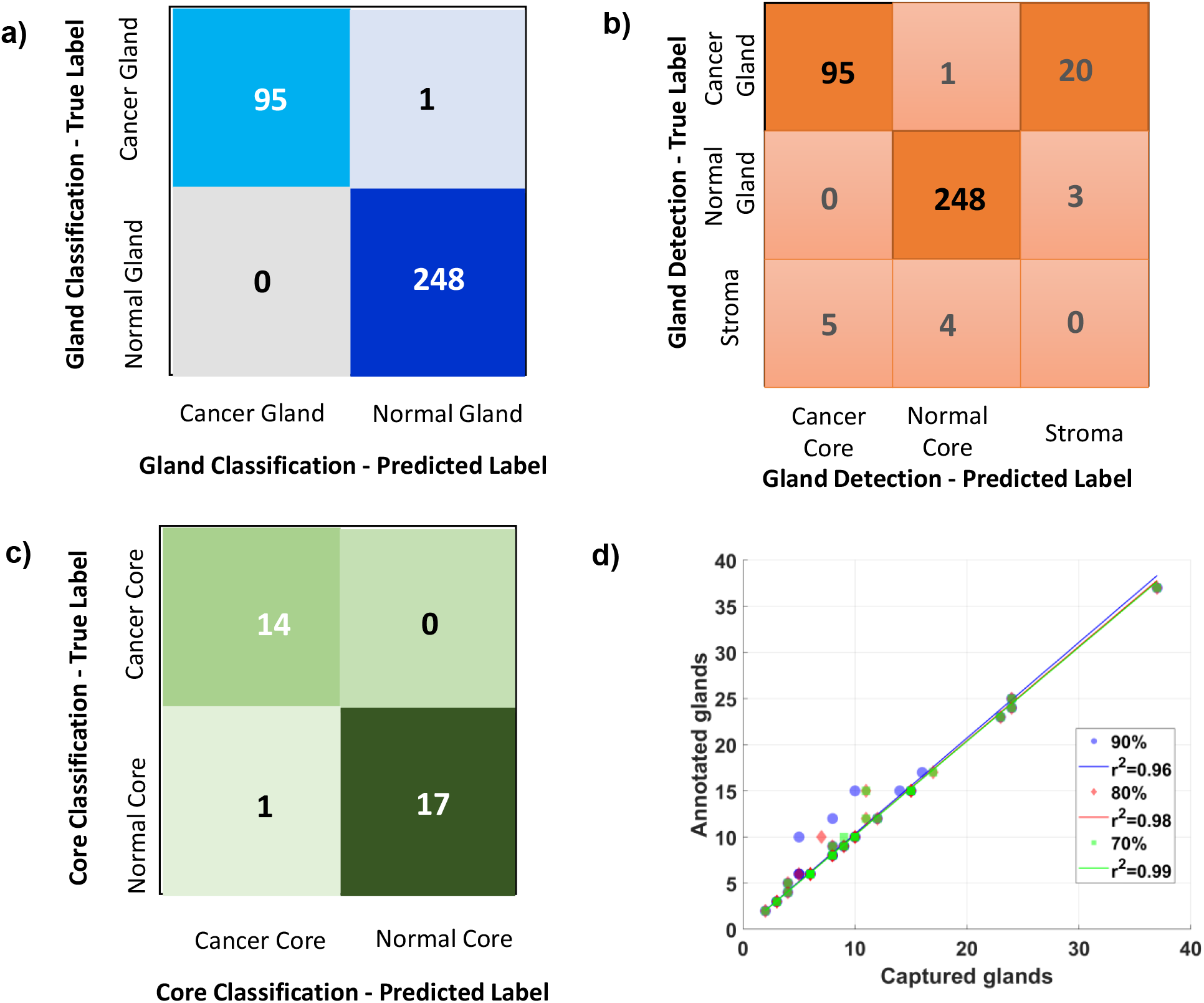
Performance of Classification, Detection, and Diagnosis. a) Confusion matrix, which shows that 95 instances of the detected cancer glands are correctly classified, while 1 is wrongly classified as normal. All the 248 instances of the detected normal glands are perfectly classified. b) Confusion matrix, which shows that 96 cancer glands are detected, with 95 correctly classified and 1 wrongly classified; 20 cancer glands are missed. 248 normal glands are detected and correctly classified, while 3 normal glands are missed. c) Confusion matrix, which shows that all the 14 cancer cores are correctly diagnosed as cancer; 17 out of the 18 normal images are correctly diagnosed, while 1 normal core is wrongly diagnosed as cancer. d) Gland detection performance at three different detection confidence scores: 90%, 80%, and 70%, as indicated.

### Overall core classification

In our research, each core contains a different number of glands, and a threshold is established for the final classification of cores. If 90%+ of the glands are classified as cancer, this image is diagnosed as cancer. If 90%+ of the glands are classified as normal, this image is diagnosed as normal. Figure 5c confusion matrix shows the final diagnosis performance for the test dataset. As ground truth, 14 images are cancer, and 18 are normal. As shown by 1^st^ row, all the 14 cancer images are correctly diagnosed; as shown by 2^nd^ row, 17 of the 18 normal images are correctly diagnosed while one is misdiagnosed as cancer.

### Detection performance at three different confidence scores

Figure 5d shows the network’s performance at three respective gland detection confidence scores, and all these three scores are generated from the 390^th^ training epoch model. At the 90% detection confidence score, 96% of all the ground truth glands are detected; at the 80% detection confidence score, 98% of all the ground truth glands are detected; at the 70% detection confidence score, 99% of all the ground truth glands are detected. Of these three scores, the 90% is chosen for the final predictions.

### Accuracy reports in classification, detecting and diagnosis

Tables 1-3 are accuracy reports, respectively, for detection, classification, and diagnosis. In Gland Detection Report (Table 1), the precision score and recall score are respectively 0.95 and 0.82 in terms of cancer gland detection, and 0.98 and 0.99 in terms of normal gland detecting. The accuracy in terms of overall gland detection is 0.91%. In Gland Classification Report (Table 2), the overall classification accuracy for all the 344 glands that are detected is 99.71%. The precision score and recall score are respectively 1.00 and 0.99 in in terms of cancer gland classification. In terms of normal gland classification, the precision score and the recall score are both perfect. In Core Classification Report (Table 3), the precision and recall scores are respectively 93% and 100% in terms of cancer core classification, and respectively 100% and 94% in terms of normal core classification; and the overall accuracy in terms of whole core classification is 97%.

**Table 1:** Gland Detection Report.

|  | **Precision** | **Recall** | **F1-score** | **Support** |
| --- | --- | --- | --- | --- |
| **Cancer Gland** | 0.95 | 0.82 | 0.88 | 116 |
| **Normal Gland** | 0.98 | 0.99 | 0.98 | 251 |
| **Stroma** | 0.00 | 0.00 | 0.00 | 9 |
| **Accuracy** |  |  | 0.91 | 376 |
| **Macro Avg** | 0.64 | 0.60 | 0.62 | 376 |
| **Weighted Avg** | 0.95 | 0.91 | 0.93 | 376 |

**Table 2:** Gland Classification Report.

|  | **Precision** | **Recall** | **F1-score** | **Support** |
| --- | --- | --- | --- | --- |
| **Cancer gland** | 1.00 | 0.99 | 0.99 | 96 |
| **Normal gland** | 1.00 | 1.00 | 1.00 | 248 |
| **Accuracy** |  |  | 1.00 | 344 |
| **Macro Avg** | 1.00 | 0.99 | 1.00 | 344 |
| **Weighted Avg** | 1.00 | 1.00 | 1.00 | 344 |

**Table 3:** Core Classification Report.

|  | **Precision** | **Recall** | **F1-score** | **Support** |
| --- | --- | --- | --- | --- |
| **Cancer** | 0.93 | 1.00 | 0.97 | 14 |
| **Normal** | 1.00 | 0.94 | 0.97 | 18 |
| **Accuracy** |  |  | 0.97 | 32 |
| **Macro Avg** | 0.97 | 0.97 | 0.97 | 32 |
| **Weighted Avg** | 0.97 | 0.97 | 0.97 | 32 |

**Table 4:**
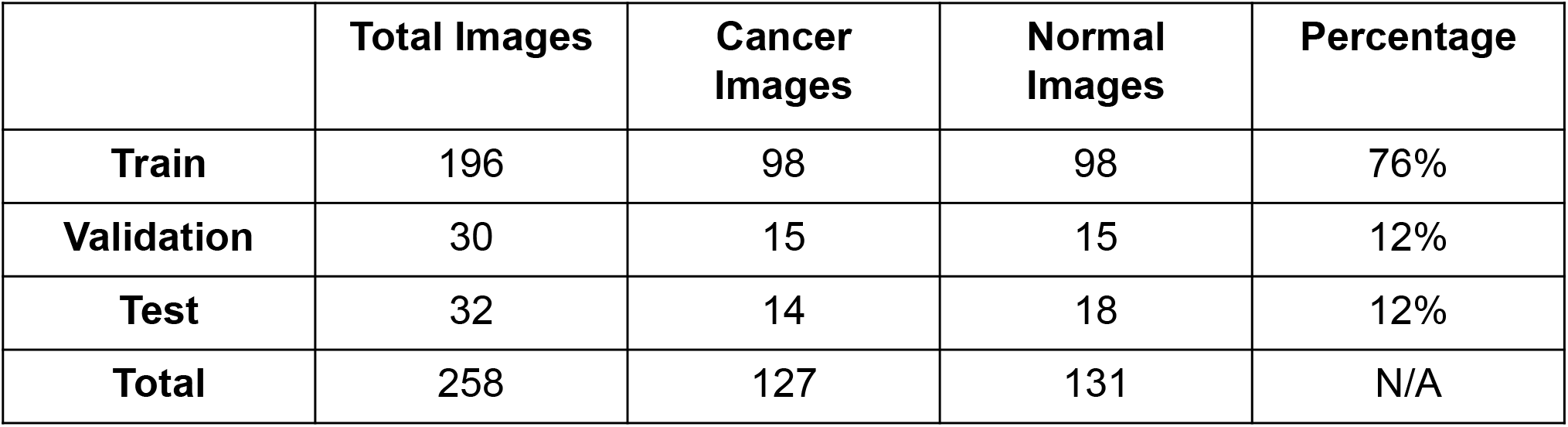
Training, Validation and Test Datasets.

### Detection performance at three different epochs

The network was trained for 500 epochs, of which the first 50 epochs trained only the RPN, classification and mask heads of the network while the remaining 450 epochs trained all the layers of the network. Three epochs were selected to show that the performance improved slowly from the 50^th^ to the 390^th^ epoch, as illustrated by Fig. 6. At the 50^th^ epoch, the network detects 87% of all the ground truth glands. At the 100^th^ epoch, the network detects 90% of the all the ground truth glands. At the 390^th^ epoch, the network detects 91% of all the ground truth glands. The improvement pace, though slow, still played a crucial role in core classification, because one error in gland detection or classification might mean an error in core classification.

**Figure 6:**
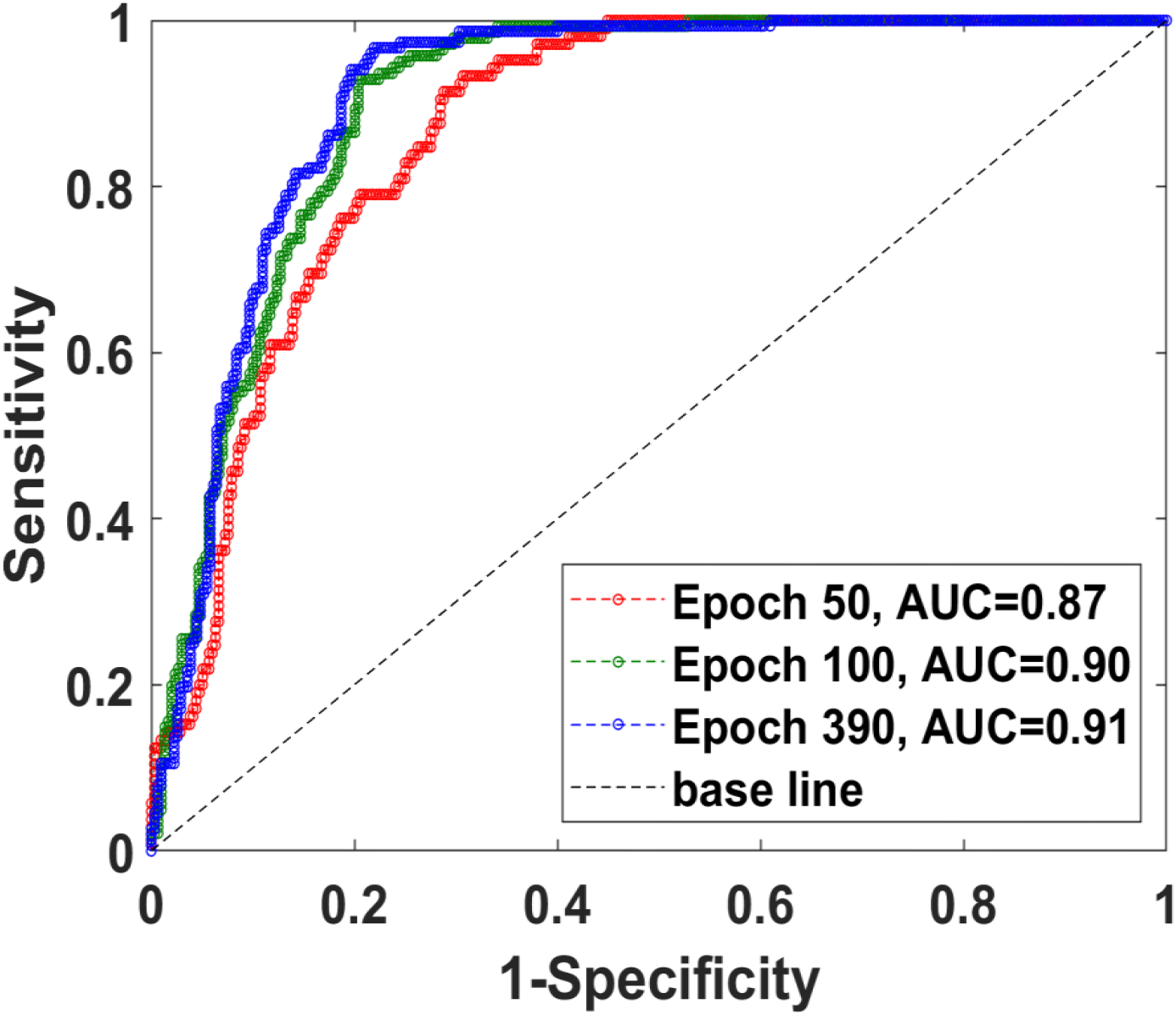
Gland Capturing Performances at Three Training Epochs. The network’s gland classification performance is shown at three different training epochs, the 50^th^, 100^th^, and 390^th^, as indicated. The AUC (Area under the ROC Curve) is 0.87, 0.90, and 0.91, respectively.

## SUMMARY AND DISCUSSION

In summary, we demonstrated promising results in colorectal tissue segmentation, classification, and whole core diagnosis, by combining Mask R-CNN deep learning network to SLIM images. The 91% of gland detection accuracy, the near perfect classification accuracy, and the 97% of whole core diagnosis accuracy show that this method can effectively assist pathologists to screen colorectal cancers. Histopathology combined with colonoscopy tissue resection remains the gold-standard for colorectal cancer diagnosis. However, we expect our method to complement valuable pathological information that can improve screening accuracy, reduce manual work, and multiply throughput at clinics. The SLIM module can be integrated with incumbent microscopes across clinics and then used a valuable tool to optimize colorectal screening workflows. Moreover, the SLIM module can also be used to quantify the aggressiveness of the cancer disease[22] or detect other types of cancer.

With its common-path interferometric geometry, SLIM is highly reliable in providing nanoscale information about tissue architecture, enabling a high sensitivity and specificity for pathological examination. The AI inference method can be integrated with SLIM acquisition software [51]. As the inference runs faster than the acquisition of a SLIM frame and it can also be operated parallelly, we anticipate performing image acquisition and diagnosis in a real-time way. In terms of overall throughput, the SLIM tissue scanner can match commercial whole slide scanners that only conduct bright field imaging on stained tissue sections.[11] Theoretically, the diagnosis, with all critical areas of interest being highlighted for pathologists, can be given while the scanning is being done.

## Data Availability

The tissue collection for this manuscript was performed in accordance with the procedures approved by the Institutional Review Board at UIC (IRB Protocol No. 2004-0317) .

## DISCLOSURES

G.P. has a financial interest in Phi Optics, Inc., a company that commercializes quantitative phase imaging technology for materials and life science applications.

## ETHNICAL STATEMENT

The tissue collection for this manuscript was performed in accordance with the procedures approved by the Institutional Review Board at UIC (IRB Protocol No. 2004-0317). The studies made for this manuscript followed the protocols outlined in the procedures approved by the Institutional Review Board at the University of Illinois at Urbana-Champaign (IRB Protocol No. 13900).

## ACKNOWLEDGMENTS

We would like to thank Mikhail Kandel for developing the software, Shamira Sridharan for imaging and annotating, and Andre Balla for diagnosing the tissues that were used for this work. This work was funded by the National Science Foundation (Grant Nos. 0939511, R01 GM129709, R01 CA238191, and R43GM133280-01). For further information, visit our website light.ece.illinois.edu.

## DATA AVAILABILITY

The data that supports the findings of this study are available from the corresponding author upon reasonable request.

## CONFLICTS OF INTEREST

The authors declare no conflicts of interest.

## Notes

### Author Declarations

The tissue collection for this manuscript was performed in accordance with the procedures approved by the Institutional Review Board at UIC (IRB Protocol No. 2004-0317) . The studies made for this manuscript followed the protocols outlined in the procedures approved by the Institutional Review Board at the University of Illinois at Urbana-Champaign (IRB Protocol No. 13900).

